# SARS-CoV-2 prevalence and transmission in swimming activities: results from a retrospective cohort study

**DOI:** 10.1101/2021.03.19.21253351

**Authors:** Martin Brink Termansen, Ask Vest Christiansen, Sebastian Frische

**Affiliations:** Department of Biomedicine, Aarhus University, Denmark; Department of Public Health, Aarhus University, Denmark

**Keywords:** Survey, questionnaire, COVID-19, epidemiology, restrictions, sports, recreational water, swimming pool

## Abstract

There is an urgent need for research on the epidemiology of severe acute respiratory syndrome coronavirus 2 (SARS-CoV-2) causing corona virus disease 2019 (COVID-19), as the transmissibility differs between settings and populations. Here we report on a questionnaire-based retrospective cohort study of the prevalence and transmission of SARS-CoV-2 among participants in swimming activities in Denmark in the last five months of 2020 during the COVID-19 pandemic.

Eight of 162 swimming activities with a SARS-CoV-2 positive participant led to transmission to 23 other participants. Overall, the percentage of transmission episodes was 4.9% (competitive swimming 8.9%; recreational swimming 1.3%). Overall, the incidence rate of transmission was 19.5 participants per 100.000 pool activity hours (corresponding values: 43.5 and 4.7 for competitive and recreational swimming, respectively).

Compliance with precautionary restrictions was highest regarding hand hygiene (98.1%) and lowest in distancing personal sports bags (69.9%). Due to low statistical power, the study showed no significant effect of restrictions.

Insight into the risk of transmission of SARS-CoV-2 during indoor swimming is needed to estimate the efficiency of restrictive measures on this and other sports and leisure activities. Only when we know how the virus spreads through various settings, optimal strategies to handle the COVID-19 pandemic can be developed.

## INTRODUCTION

Corona virus disease 2019 (COVID-19) has since its outbreak in December 2019 caused millions of deaths worldwide and is declared a pandemic by the World Health Organization[1]. The number of new confirmed cases was by January 2021 falling for the first time since the outbreak, indicating better disease control and posing discussion of ending lockdowns and curfews[2]. COVID-19 is caused by the newly emerged severe acute respiratory syndrome coronavirus 2 (SARS-CoV-2; family *Coronaviridae*, genus *Betacoronavirus*)[3, 4]. SARS-CoV-2 is recognized to be transmitted primarily by droplets and close contact and secondarily by fomites and aerosols and possibly fecal-oral, mother-to-child and bloodborne transmission[5].

To limit the spread of SARS-CoV-2, the Danish Government mandated an extensive lockdown of the Danish society including all indoor sports activities on March 11, 2020[6]. On June 8, 2020, indoor sports activities were reopened but subjected to restrictive measures[7]. As regards indoor water sports “Corona Restrictions in Water Sports” (“Vandsportens Coronaregler”) was published by the Danish Swimming Federation (DSF) with guidance from the Danish health authorities[8]. The restrictions involved among other things: distancing in and out of the water; thorough sanitation and showering; limiting physical activity around the pool area; limiting group size to maximum 50 participants; and limiting use of shared equipment. Simultaneously, all swimming pools were advised by the Danish Environmental Protection Agency to raise the chlorine content to at least 0.8 mg/L from the normal average between 0.4-0.8 mg/L[9].

Due to limited knowledge and an unpredictable behavior of coronavirus, the extent of transmission of SARS-CoV-2 in swimming activities compared to other sports and leisure activities is still debated. Indoor swimming has conditions similar to other indoor activities for transmission by droplets and close contact. The main structural difference to other sports is the medium used for swimming: water. The majority of swimming pool-related disease outbreaks are caused by enteric viruses and by adenoviruses in particular[10]. As such, fecal-oral transmission is the most common cause of general viral transmission through swimming pool water.

While possible, fecal-oral transmission of SARS-CoV-2 is currently considered a minor contributor to overall disease spread[5, 10, 11]. The virus cannot replicate outside the host’s tissue. Therefore, all presence of SARS-CoV-2 in swimming pool water may be the result of direct contamination by swimmers through release of respiratory droplets or body fluids.

Viral transmission in swimming pools are generally related to low concentrations of disinfectants such as chlorine[12]. High susceptibility to chlorine seems to be applicable to human coronaviruses specifically[13]. Further, enveloped viruses are recognized to be instable in water, though this has been challenged by SARS-CoV-2 due to a particularly hard outer shell[14]. However, we are not aware of any scientific reports of SARS-CoV-2 epidemiology in swimming pools.

To fill this knowledge gap, this study was designed as a retrospective questionnaire-based cohort study to quantitatively describe the extent of transmission of SARS-CoV-2 at indoor swimming activities in Danish swimming clubs during the last five months of 2020. We aimed to quantify both the number of risk episodes, where a SARS-CoV-2 positive subject was participating in a swimming activity, and transmission episodes, where other participants in a risk episode subsequently tested positive for SARS-CoV-2.

As mentioned above, a range of restrictions on the normal use of swimming facilities was in effect in Danish swimming clubs during the study period. We therefore also aimed to investigate the extent of compliance to these restrictions and if compliance could be related to the extent of transmission of SARS-CoV-2 during swimming activities.

## MATERIALS AND METHODS

### Situational background

From June 8, 2020 to December 9, Danish swimming clubs were allowed to resume swimming activities under a set of precautionary restrictions to prevent spread of SARS-CoV-2 virus[7, 8, 15]. Overall, the clubs reached close to normal activity levels in the pools, but the number of meets and competitions were reduced and public access to swimming pools was not allowed.

The Danish public health system provided free access to polymerase chain reaction (PCR) tests for all citizens based on a self-service booking system. Timeslots for tests were normally available within 24 hours and results normally available within 48 hours after testing.

### Study design

This is a questionnaire-based, nationwide, retrospective cohort study of swimming activities in Danish swimming clubs in the last five months of 2020. Certain analyses only include data from swimming activities between August 3 and December 6, 2020 (week 32 to week 49) when all Danish swimming clubs were allowed full activity[7, 15].

### Participants

Geographically, this survey covered all five Danish regions. The sampling frame was DSF’s official list of member clubs. One official contact person from each swimming club was invited by e-mail to participate (*n* = 298). The sampling did not include swimming clubs not under the auspices of DSF. As all clubs under DSF were invited, sample size was not calculated beforehand. The exact number of swimming clubs in Denmark is not known but estimated >75% of indoor swimming clubs in Denmark are members of DSF. This includes an estimated >95% of Danish swimming club members.

### Data collection

A questionnaire in Danish language was developed and distributed electronically via SurveyXact (Rambøll Management Consulting, Aarhus, Denmark) on December 17, 2020. Deadline for replies was January 14, 2021. Two e-mails and in some cases one telephone reminder were given to non-responders in early January 2021. It took 10-30 minutes to fill in the questionnaire depending on number of reported risk episodes. No validation study of the questionnaire was performed; however, before distribution, all question-and-answer options were piloted for comprehensibility among the research group and other peers and the questionnaire was revised accordingly.

A total of 208 questions were included in the questionnaire, though the number of questions the individual respondent would meet depended on their answers. Questions were multiple choice and open-ended. Multiple choice questions were provided with “I do not know” and “Other” (open ended) options where appropriate. The questionnaire collected information about the respondent’s position in the swimming club, the swimming club and its pool premises, pool free chlorine level and pool activity hours in different categories of swimming activities. Further, the questionnaire collected the following information on specific risk episodes: category of swimming activity, number of participants, if any other participants subsequently tested positive for SARS-CoV-2, compliance to restrictions, and a free-text field for additional information.

To prevent large swimming clubs with high numbers of risk episodes from not responding due to the time it would take to answer the electronic questionnaire, a manual questionnaire recording similar information but with a less detailed approach was developed. During follow-up of non-responders, this questionnaire was used on request.

During analysis, it became clear that the formulation of one question resulted in ambiguous answers. The question was: “Were all close contacts at the risk episode subsequently tested?” To this, 13 clubs replied “Other” and specified some details, which were evaluated. Data from clubs that instructed participants to be tested according to national guidelines were included for estimation of transmission fraction. Follow-up phone calls were made to 11 swimming clubs who had replied “Do not know” to the question. Based on these phone calls, data were included from clubs who instructed participants to be tested according to national guidelines. Data from clubs answering “No” to the question were excluded and data from clubs answering “Yes” were included.

Some clubs reported risk episodes and transmission episodes not related specifically to indoor swimming activities (other sport activities, training camps etc.). These episodes were not included in the study.

### Outcome measures

Primary outcome: The fraction of risk episodes that resulted in other participants subsequently testing positive for SARS-CoV-2.

Secondary outcomes: The incidence of SARS-CoV-2 transmission during swimming activities and the relation between precautionary restrictions and SARS-CoV-2 transmission episodes.

### Data processing and statistical analysis

Data were analyzed using Microsoft Excel v. 16.43 for Mac (Microsoft Corporation, Seattle, WA) and statistical analyses were conducted with Stata/IC v. 16.1 for Mac (StataCorp LCC, College Station, TX). Figures were made using GraphPad Prism version 9.0.2 for Mac (GraphPad Software, San Diego, CA) or Lucidchart (www.lucidchart.com).

We intended to sample proportionally equal during the study period as well as geographically in the five Danish regions and assumed that the number of risk episodes observed overall and within each category of swimming activities would be proportional to the general incidence of SARS-CoV-2 virus in Denmark in the study period.

To test if our sampling was biased in time, we performed a simple linear regression to assess the ability of the weekly SARS-CoV-2 positive cases in Denmark[16] to predict the weekly number of sampled risk episodes. Further, to test any geographic bias, we compared the regional proportions of all publicly available PCR confirmed SARS-CoV-2 positive cases in Denmark[16] to the observed regional proportions of risk episodes. Hypotheses of no difference in proportions were assessed using z-tests with an alpha of 0.05. Further, we graphically compared weekly regional and national SARS-CoV-2 infection incidence to observed weekly regional and national risk episodes. 126 risk episodes for which week numbers were available and that were held in week 32-49 in 2020 (both weeks included) were included in these analyses.

Obviously, the validity of the reported data on risk episodes depended heavily on the extent of the reporting person’s knowledge of swimmers’ SARS-CoV-2 status. To identify potential risk of bias we assessed the difference in sampling of risk episodes in different swimming activity categories by comparing the number of risk episodes reported from each category per pool activity hours (hours of activity per swimming pool premise in that particular category) using the Poisson model and Fishers exact test with an alpha of 0.05.

For incidence rates the period at risk was defined as pool activity hours. Responses from swimming clubs were included if these reported both weekly pool activity hours and week numbers of any potential risk episodes. Swimming clubs only reporting risk episodes, where close contacts were not subsequently asked to be tested, were excluded. Swimming clubs reporting risk episodes both with and without testing of close contacts were included. In total, five reported transmission episodes leading to transmission to 15 other participants were included to estimate incidence rates. The period was limited to week 32-49 in 2020 (both weeks included) for competitive swimming and water polo, and week 35-49 (both weeks included; week 42 excluded) for recreational swimming. The weeks differed due to public holidays affecting recreational swimming activities only. The reported weekly pool activity hours were multiplied according to these periods. No confidence intervals were given, as the Poisson distribution was challenged on independence due to transmission to more than one individual at some risk episodes.

To describe the circumstances under which this study was done, we reported descriptive statistics of the level of compliance to nationwide restrictions and number of participants in risk episodes. The level of compliance to restrictions was reported as a bar chart showing the proportions of risk episodes following restrictions. As the reporting of risk episodes may not be representative of all swimming activities, no confidence intervals were reported for these data. The number of participants in risk episodes were presented as a scatter dot plot with marking of the mean.

112 risk episodes at which close contacts afterwards were tested for SARS-CoV-2 by PCR were included in the analysis of effect of the individual restrictions. Effect was measured as risk difference and risk ratio of transmission of SARS-CoV-2. A Fishers exact test was used to assess the hypothesis of no difference with an alpha of 0.05. Estimates were given with 95% confidence intervals and the respective power. The same 112 activities were included in the analysis of the summarized effect of restrictions. We made a graphical presentation of the number of episodes that followed between 1 and 10 restrictions for all risk episodes and for transmission episodes.

## RESULTS

We received 183 replies to the electronic questionnaire and three replies to the manual questionnaire (Figure 1a). Of these 11 were excluded as duplicates, and three were excluded due to deficient or conflicting answers that could not be corrected despite several e-mail exchanges. This led to inclusion of answers from 172 of the 298 invited swimming clubs (response rate: 57.7%). The sampled swimming clubs had in total 159,807 members, corresponding to 82.7% of all members in DSF member clubs (data from DSF). The respondents often served multiple roles in the swimming clubs, as 112 of respondents (65.1%) were board members, 55 (32.0%) were leaders, 40 (23.3%) were trainers, 27 (15.7%) were instructors and 20 (11.6%) were swimmers.

**Figure 1.**
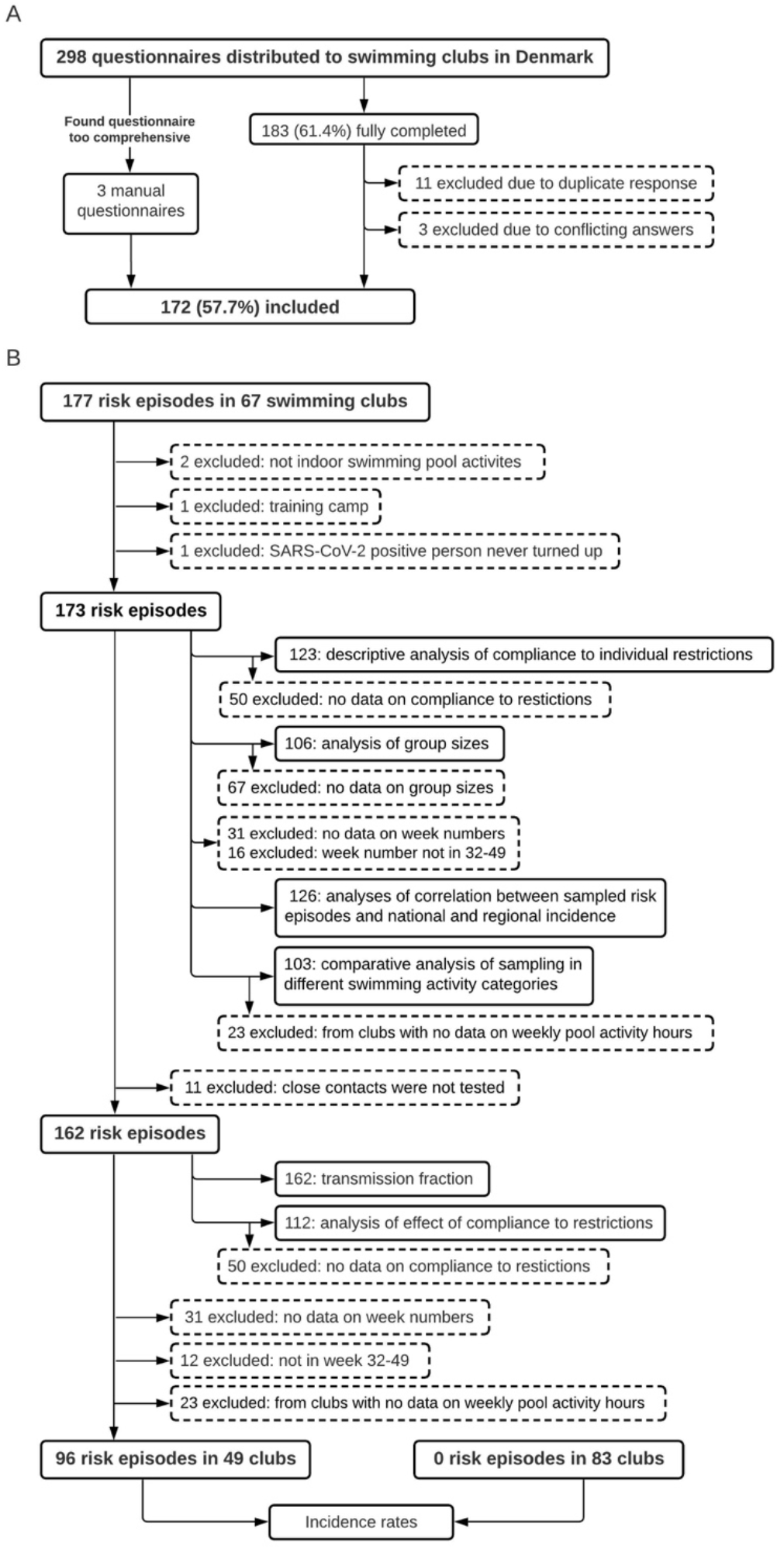
Inclusion and exclusion of respondents and answers. A. Three manual questionnaires were on request sent by e-mail to three larger swimming clubs to get a more efficient overview of their many risk episodes (swimming activity where a SARS-CoV-2 positive subject was participating). Three online responses were excluded due to inconsistency such as different answers on number of risk episodes or lack of knowledge on central questions, i.e. number of risk episodes. B. After removal of reported risk episodes not solely associated with indoor swimming activities, 173 risk episodes from 66 swimming clubs were available for the analyses. The remaining 106 swimming clubs reported zero risk episodes. The questionnaire was built to retrieve detailed information on up to four risk episodes (risk episodes above four was followed up by e-mail and phone), and the respondents were always given the opportunity to answer “I do not know”. However, all details were not reported for all risk episodes, so only a subset of risk episodes could be included in each analysis, e.g. only 123 risk episodes reported data on compliance to restrictions. 11 risk episodes were excluded in primary and secondary outcome analyses due to no testing of close contacts.

All 172 swimming clubs reported if they had experienced any risk episodes, and whether these had led to transmission of SARS-CoV-2 to other participants. 179 risk episodes were reported in 67 swimming clubs (Figure 1b). Only indoor swimming activities were included. No training camps were included. This led to inclusion of 173 risk episodes in 66 swimming clubs. 162 risk episodes in 63 swimming clubs were included in analyses on fractions or risks of transmission, as 11 risk episodes were excluded due to lack of testing of close contacts. None of these 11 risk episodes were reported to lead to transmission of SARS-CoV-2. Complete information could not be obtained on all questions for all risk episodes. 132 swimming clubs, that had not only had risk episodes without testing of close contacts, and that reported week number of potential risk episodes, reported weekly pool activity hours in week 32-49 in 2020 (Table 1).

**Table 1.** **Weekly pool activity hours in 132 Danish swimming clubs in week 32-49 in 2020**.

|  | Total | Competitive swimming | Recreational swimming<sup>§</sup> | Water polo |
| --- | --- | --- | --- | --- |
| Pool activity hours<sup>†</sup> | 4,915.37<sup>‡</sup> | 1,658.46 | 3,066.57 | 222.50 |

<sup>†</sup>Number of weekly hours the swimming pool premises were used for this swimming activity category.
<sup>‡</sup>As different swimming activity categories may take place simultaneously in the same swimming pool premises, the total does not sum up the three disciplines.
<sup>§</sup>All activities excluding competitive swimming and water polo. Including swimming lessons, aqua fitness, water gymnastics, warm water exercise, aquaphobia training and family swimming.
Responses from 132 swimming clubs, as these reported pool activity hours, reported week numbers of potential risk episodes, and not all risk episodes had no testing of close contacts.

### Validity of reported data

Weekly national SARS-CoV-2 incidence and national number of sampled risk episodes were correlated over time with and *R*^*2*^ of 0.814 and the linear regression was statistically significant (F(1,16) = 69.77, *p* < 0.0001) (Figure 2).

**Figure 2.**
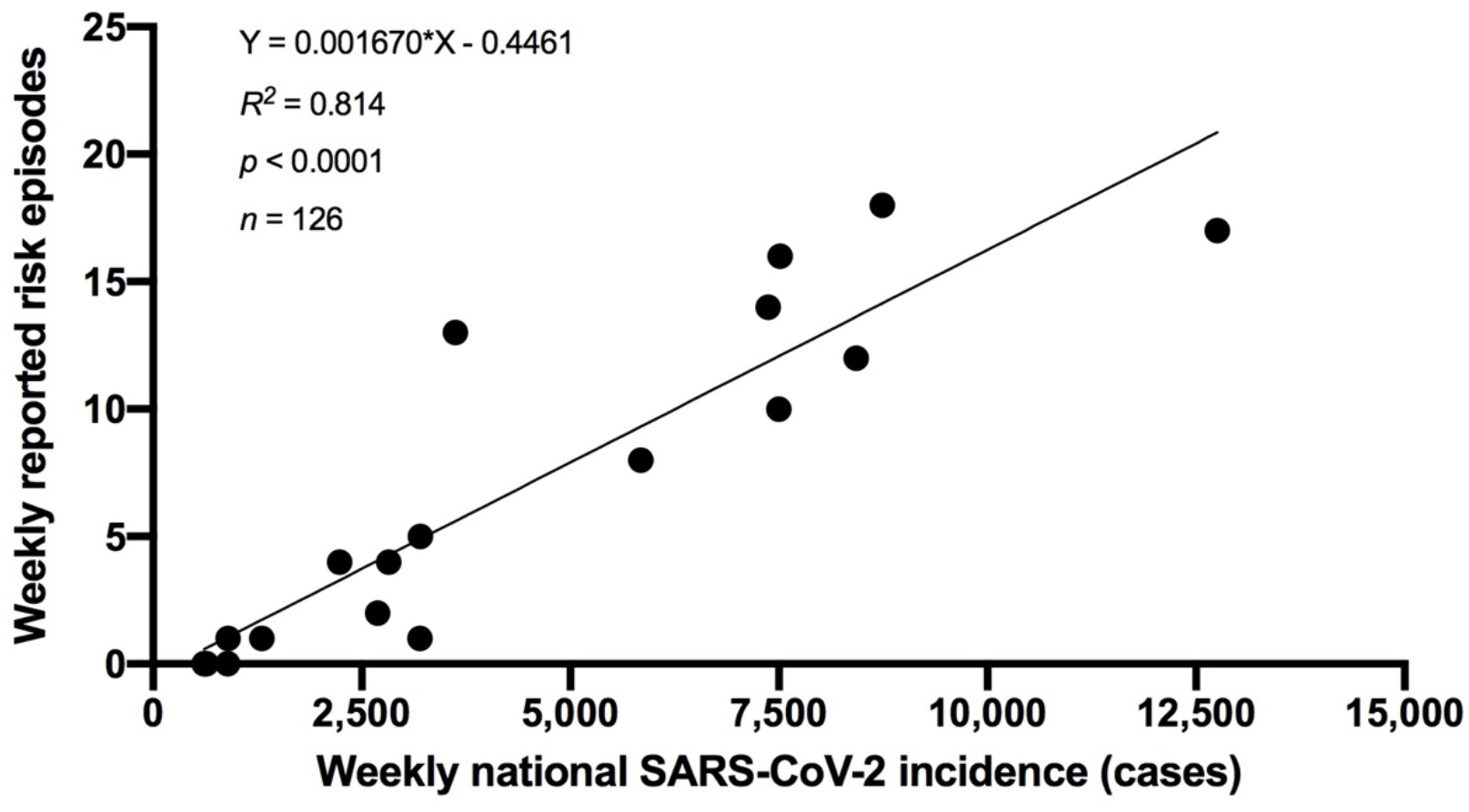
Linear correlation between weekly national SARS-CoV-2 infection incidence and number of weekly reported risk episodes. Black dots are weekly reported risk episodes for all swimming activity categories in week 32-49 (*n* = 126). Total number of weekly new SARS-CoV-2 positive cases in Denmark were diagnosed by polymerase chain reaction-based tests, which was free and easily available at the time (data from covid19.ssi.dk).

There was no statistically significant difference in regional proportions of SARS-CoV-2 positive cases and regional proportions of risk episodes (Table 3). The weekly distribution of 126 risk episodes to the regional and national incidence is shown in Figure 3.

**Figure 3.**
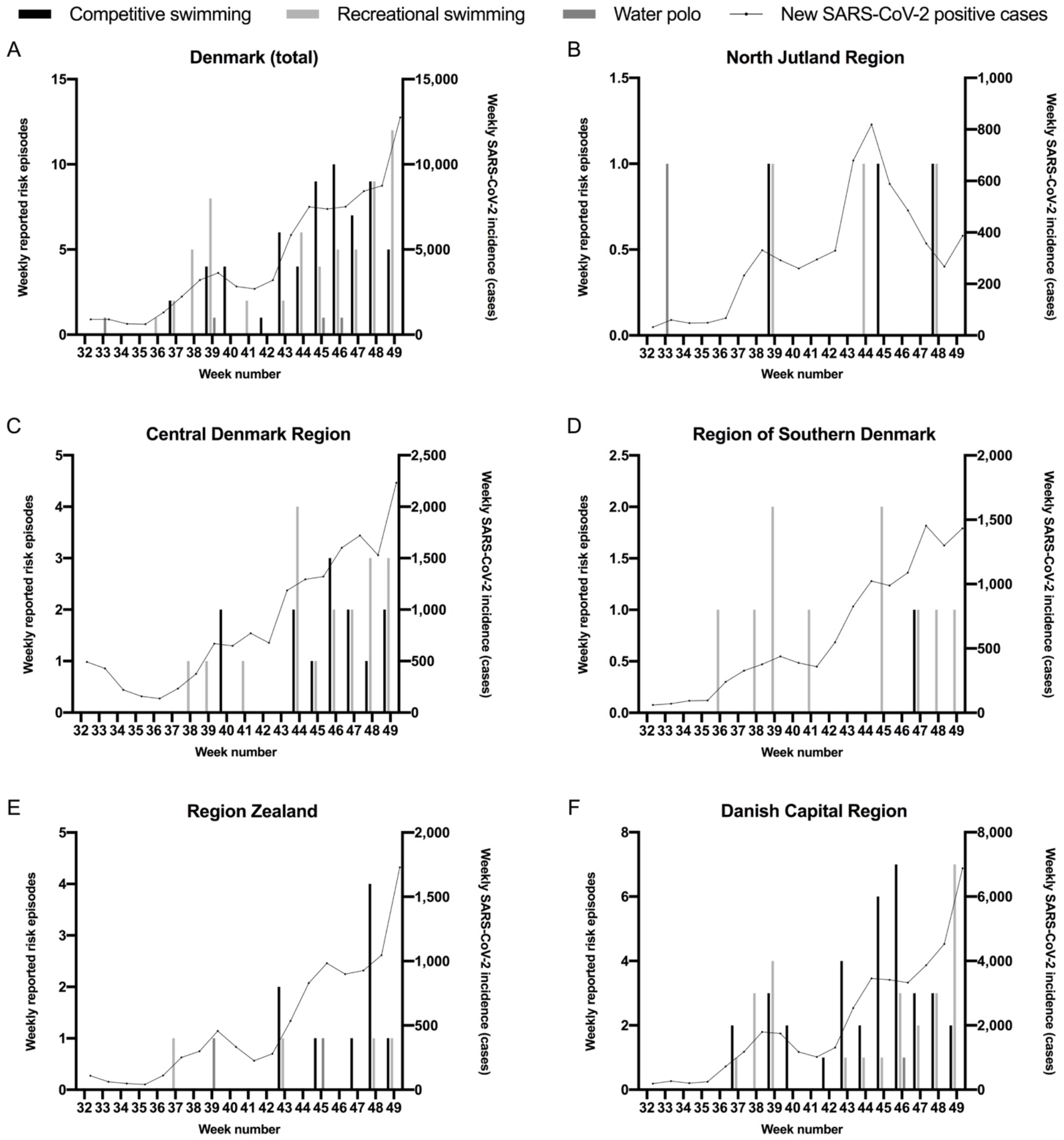
Geographic distribution of weekly reported risk episodes compared to total weekly SARS-CoV-2 infection incidence. 126 risk episodes (swimming activities where a SARS-CoV-2 positive subject was participating) were included, as they were reported to happen in week 32-49 in 2020. Total number of weekly new SARS-CoV-2 positive cases from each geographic region were diagnosed by polymerase chain reaction-based tests, which was free and easily available at the time (data from covid19.ssi.dk).

A higher number of risk episodes per pool activity hours was seen in competitive swimming (153.1 risk episodes per 100.000 hours) compared to recreational swimming (115.4 risk episodes per 100.000 hours), though this was not statistically significant (*p* = 0.1621). There was no statistically significant difference in rate of risk episodes between water polo (99.9 risk episodes per 100.000 hours) and recreational swimming (*p* = 1.000) or water polo and competitive swimming (*p* = 0.870). 103 risk episodes in week 32-49 from swimming clubs reporting pool activity hours were included in this analysis.

### Fraction of risk episodes resulting in transmission

Of 162 total risk episodes eight (fraction: 4.9%) led to transmission to 23 participants (Table 2). In competitive swimming seven of 79 risk episodes (fraction: 8.9%) led to transmission to 21 participants. In recreational swimming one of 79 risk episodes (fraction: 1.3%) led to transmission to two participants. In water polo there was reported no transmission.

**Table 2.** Descriptive results for each category of swimming activity in 172 swimming clubs.

|  | Total | Competitive swimming |  | Recreational swimming<sup>□</sup> |  | Water polo |  |
| --- | --- | --- | --- | --- | --- | --- | --- |
|  |  | Training | Competition | Training | Competition | Training | Competition |
| Risk episodes<sup>†</sup> | 162 | 69 | 10 | 79 | 0 | 3 | 1 |
| Transmission episodes<sup>‡</sup> | 8 | 5 | 2 | 1 | 0 | 0 | 0 |
| Fraction<sup>§</sup> | 0.049 | 0.072 | 0.200 | 0.013 | - | 0.000 | 0.000 |
| Participants infected<sup>¶</sup> | 23 | 15 | 6 | 2 | 0 | 0 | 0 |

<sup>†</sup>Swimming activities where a SARS-CoV-2 positive subject was participating. Only risk episodes where all close contacts afterwards were tested were included.
<sup>‡</sup>Risk episodes where other participants subsequently tested positive for SARS-CoV-2.
<sup>§</sup>Transmission episodes divided by risk episodes.
<sup>¶</sup>Number of infected participants other than the initially infected person.
<sup>□</sup>All activities excluding competitive swimming and water polo. Including swimming lessons, aqua fitness, water gymnastics, warm water exercise, aquaphobia training and family swimming.

**Table 3.** **Comparison of regional proportions of reported risk episodes in Danish swimming clubs to regional proportions of nationwide Danish SARS-CoV-2 infection incidence**.

| Danish region | Regional SARS-CoV-2 cases<sup>†</sup> | Proportion of national cases<sup>†</sup> | Risk episodes<sup>‡</sup> in the region | Proportion of national risk episodes | P value<sup>§</sup> |
| --- | --- | --- | --- | --- | --- |
| North Jutland | 5578 | 0.070 | 7 | 0.056 | 0.519 |
| Central Denmark | 15693 | 0.198 | 31 | 0.246 | 0.173 |
| Southern Denmark | 11109 | 0.140 | 11 | 0.087 | 0.089 |
| Zealand | 9172 | 0.115 | 15 | 0.119 | 0.901 |
| Danish Capital | 37870 | 0.477 | 62 | 0.492 | 0.732 |

<sup>†</sup>Data from covid19.ssi.dk (downloaded 19 February 2021).
<sup>‡</sup>Swimming activities where a SARS-CoV-2 positive subject was participating.
<sup>§</sup>Z-test. Significance level = 0.05.
SARS-CoV-2 infection incidence and risk episodes were only included for week 32-49 in 2020 (including both weeks).

### Incidence rates of SARS-CoV-2 transmission

Overall, swimming activities in the defined period resulted in transmission of SARS-CoV-2 to 19.5 participants per 100,000 pool activity hours.

In competitive swimming the period at risk for week 32-49 (both weeks included) were estimated to 29,852 pool activity hours. In this period competitive swimming resulted in transmission of SARS-CoV-2 to 43.5 participants per 100,000 pool activity hours. In recreational swimming the period at risk for week 35-49 (both weeks included; week 42 excluded) were estimated to 42,932 pool activity hours. In this period recreational swimming resulted in transmission of SARS-CoV-2 to 4.7 participants per 100,000 pool activity hours. In water polo the period at risk for week 32-49 (both weeks included) were estimated to 4,005 pool activity hours. As no transmission episodes were recorded, no incidence rate for this category was calculated.

### Compliance to restrictions and their effect

For 123 risk episodes, 61 swimming clubs reported the compliance to each of 10 specific restrictions (Figure 4). The highest compliance was to washing hands upon arrival and at departure from the swimming pool premises (98.1%). The lowest compliance was to keeping distance between personal bags, equipment etc. (69.9%).

**Figure 4.**
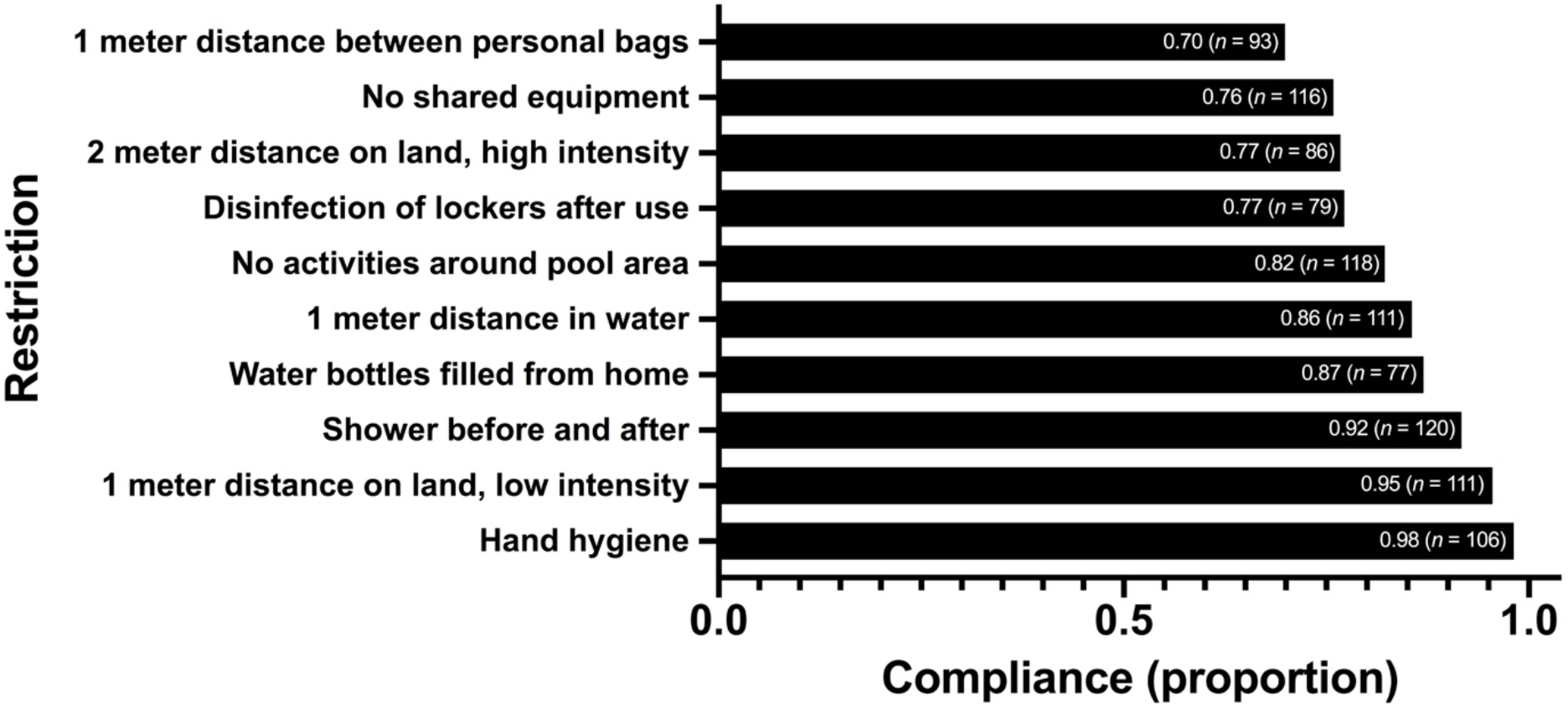
Compliance to specific individual restrictions. Fractions of 123 risk episodes (swimming activities where a SARS-CoV-2 positive subject was participating) in 61 Danish swimming clubs that followed each specific restriction during the COVID-19 pandemic in 2020. Episodes included both competitive swimming, recreational swimming and water polo. Number of responses to questions on individual restrictions varied due to the respondent’s knowledge.

For 112 risk episodes, the swimming clubs reported compliance to each of 10 specific restrictions, and all participants were afterwards asked to be tested for SARS-CoV-2, which enables assessment of transmission at the episode. From this sample, all restrictions did not change the risk of transmission by statistical significance (Table 4). However, for the four restrictions that had transmission episodes in both groups, it was possible to calculate power, which proved to be low (power < 0.25). The effect of hand hygiene was not possible to estimate because all risk episodes of this analysis complied to hand hygiene.

**Table 4.** Effect of every individual restriction.

| Restriction | RD<sup>‡</sup> (95% CI) | RR<sup>§</sup> (95% CI) | *P* value<sup>†</sup> | Power | *n* (f/nf)<sup>¶</sup> |
| --- | --- | --- | --- | --- | --- |
| 1 meter distance between sports bags | -7.9 (-24.1;8.2) | 0.44 (0.11;1.83) | 0.359 | 0.120 | 84 (63/21) |
| No shared equipment | 9.8 (3.3;16.2) | - | 0.194 | - | 106 (82/24) |
| 2 meters distance on land, high intensity | 12.5 (4.4;20.6) | - | 0.341 | - | 79 (64/15) |
| Disinfection of lockers after use | 7.4 (0.4;14.4) | - | 0.570 | - | 69 (54/15) |
| No physical activities around pool area | -10.4 (-26.7;5.9) | 0.31 (0.07;1.26) | 0.119 | 0.247 | 107 (87/20) |
| 1 meter distance in water | 6.8 (1.6;12.1) | - | 1.000 | - | 101 (88/13) |
| Water bottles filled from home | -15.2 (-40.5;10.2) | 0.24 (0.05;1.27) | 0.139 | 0.212 | 72 (62/10) |
| Shower before and after | -4.1 (-25.2;17.0) | 0.63 (0.09;4.57) | 0.510 | 0.029 | 109 (100/9) |
| 1 meter distance on land, low intensity | 5.1 (0.7;9.5) | - | 1.000 | - | 101 (98/3) |
| Hand hygiene | - | - | - | - | 96 (96/0) |

<sup>†</sup>Fisher's exact test. Significance level = 0.05.
<sup>‡</sup>RD = risk difference. %-point.
<sup>§</sup>RR = risk ratio.
<sup>¶</sup>f = number of responses reporting compliance. nf = number of responses reporting non-compliance. *n* = total number of responses.
Total number of responses to questions on individual restrictions varied due to whether the respondent knew the answer or not. Risk refer to the probability of transmission of SARS-CoV-2 at a swimming activity, where a SARS-CoV-2 positive subject was participating. Empty cells (hyphens) are due to a zero in one or more cells of the 2x2 contingency table. All respondents answering the question on hand hygiene reported good compliance, why no RD or RR were estimated for this restriction.

Although numbers are small, the number of restrictions that were followed at all 112 risk episodes and at the eight transmission episodes, does not appear to differ (Figure 5).

**Figure 5.**
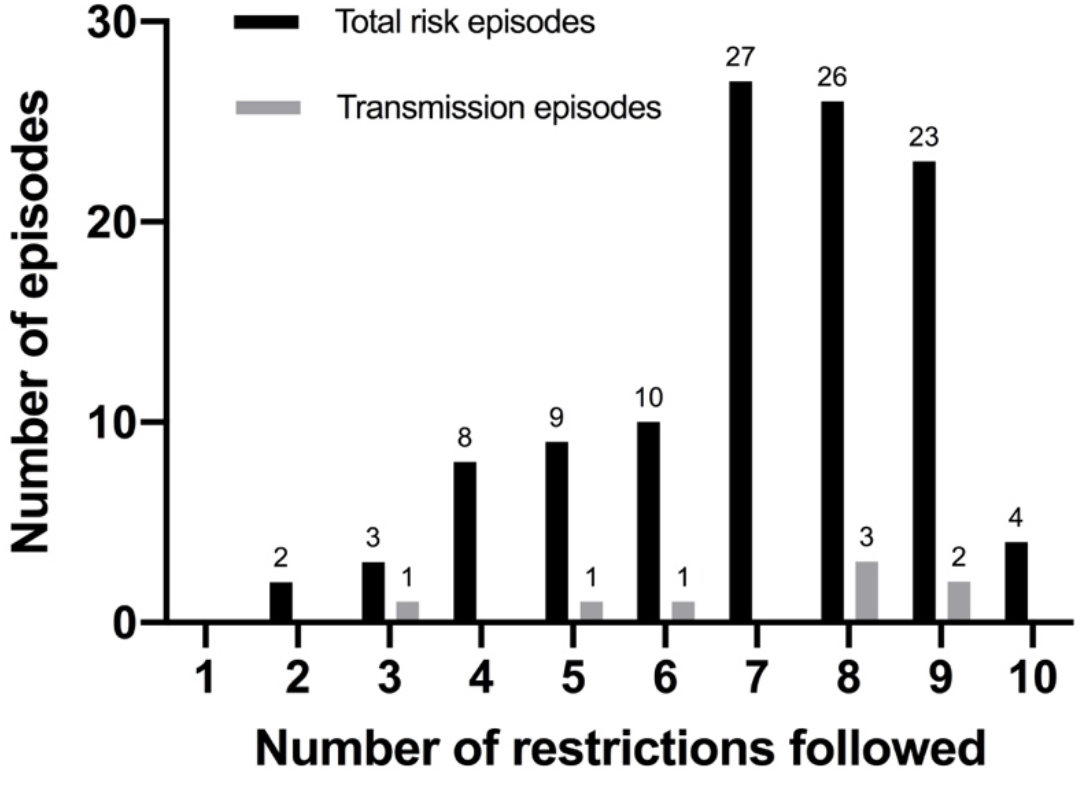
Number of restrictions followed at risk and transmission episodes. Number of specific restrictions reported to have been followed at 112 risk episodes at which transmission could have been detected if present. Risk episodes are swimming activities where a SARS-CoV-2 positive subject was participating. Transmission episodes are risk episodes where other participants subsequently tested positive for SARS-CoV-2.

For 106 risk episodes, 55 swimming clubs reported the group size (Figure 6). The mean group size at these risk episodes was 14.6 (95% CI: 12.9;16.3). The restriction of group sizes to 50 participants, including trainers, was adhered to at all but one risk episode.

**Figure 6.**
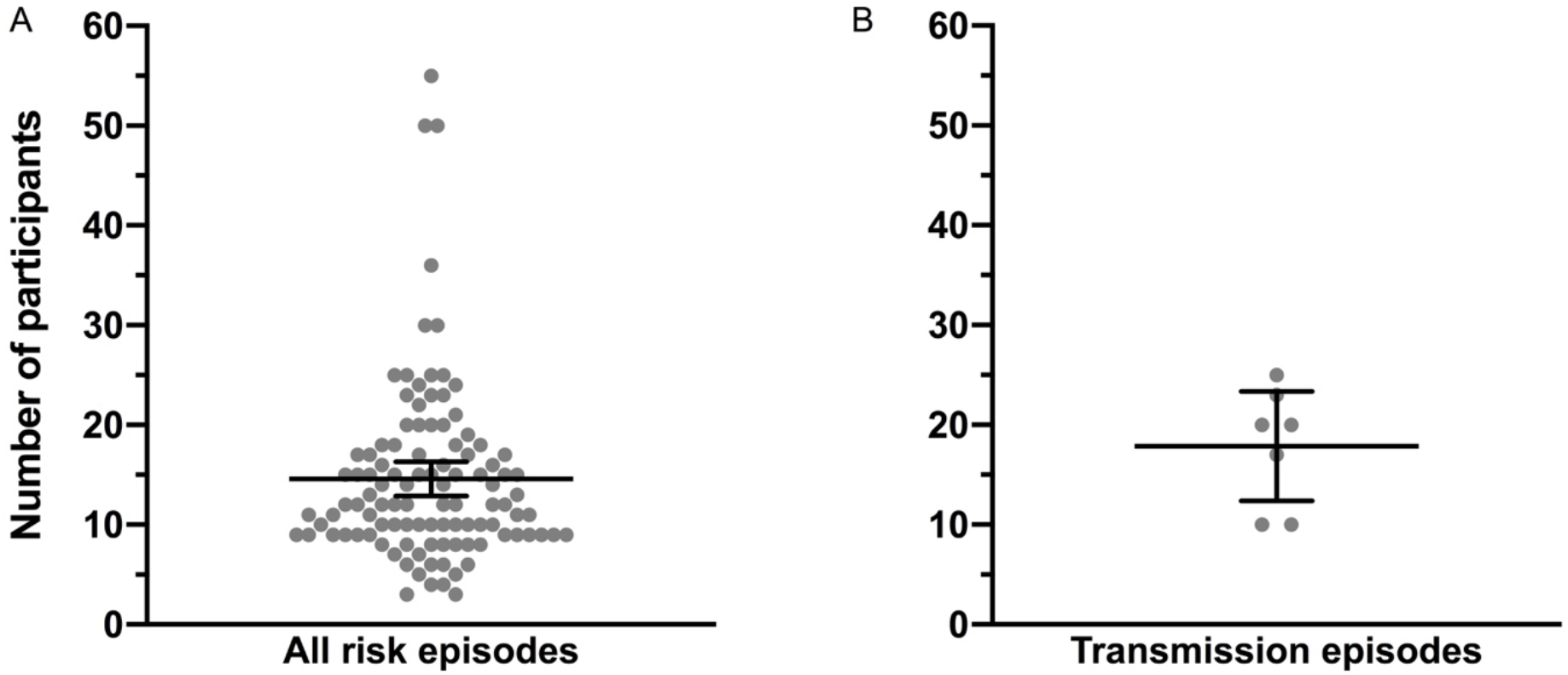
Group size at risk and transmission episodes. A. 106 risk episodes (swimming activities where a SARS-CoV-2 positive subject was participating) in 55 swimming clubs. Minimum: 3. Maximum: 55 Mean: 14.58 participants. 95% confidence interval: (12.87;16.39). Transmission episodes are risk episodes where other participants subsequently tested positive for SARS-CoV-2. These seven episodes are also included in Figure 6A. Minimum: 10. Maximum: 25. Mean: 17.86. 95% confidence interval: (12.37;23.34). Y-axis shows total number of participants in the groups of the SARS-CoV-2 positive subject. More people might have been in the swimming pool premises but keeping two meters distance to other groups. Every grey dot represents one group at one risk episode. Wide horizontal bar represents mean, narrow horizontal bars represent 95% confidence limits.

### Chlorine concentrations

A low response rate was seen on free chlorine concentrations in swimming pools due to lack of knowledge on this subject. This is likely due to the fact that chlorine concentrations are not the responsibility of most swimming clubs, but instead the local municipality that administers swimming pools.

34 respondents (19.8%) knew their swimming pools’ free chlorine concentrations. Of these 16 clubs (43.2%) reported an increase in free chlorine concentrations at least once after the beginning of the COVID-19 pandemic. Before the COVID-19 pandemic the average swimming pool free chlorine concentration level was reported to be between 0.5-0.8 mg/L; after the increases the average level was reported to be between 0.8-1.2 mg/L. No reports of lowered free chlorine concentrations were recorded.

## DISCUSSION

This study reports 162 risk episodes including eight transmission episodes at which SARS-CoV-2 is likely to have been transmitted during indoor swimming activities. These numbers should be considered in the perspective, that the survey is based on the activity of 82.7% of all members of DSF member clubs (159,807 persons). In addition, the prevalence of SARS-CoV-2 in Denmark as well as the restrictive measures in effect in the study period should be kept in mind.

The percentage of risk episodes leading to transmission of SARS-CoV-2 is overall 4.9%, and although this percentage appears to differ between competitive swimming (8.9%) and recreational swimming (1.3%), there was no statistically significant difference (Fisher’s exact test: *p* = 0.063). The numbers are estimates of the risk of transmission to occur at an indoor swimming event if a participant is SARS-CoV-2 positive. Due to methodological limitations, the apparent difference between competitive swimming and recreational swimming should be interpreted with caution (see limitations below).

As the risk episodes exposed on average 13.6 (95% CI: 11.9;15.3) participants to SARS-CoV-2, in total 2,201 (95% CI: 1,923;2,479) swimmers are estimated to have been exposed to SARS-CoV-2 at the recorded risk episodes. Of these swimmers, 23 subsequently tested positive for SARS-CoV-2, leading to an estimated risk of infection of 1.0 (95% CI: 0.9;1.2)% by participating in a risk episode. This number is an estimate of a participant’s risk of getting infected with SARS-CoV-2 at an indoor swimming activity (under the restrictions prevailing in the study period) if another participant is SARS-CoV-2 positive. For comparison, a cohort study showed SARS-CoV-2 transmission from non-hospitalized COVID-19 patients to 12-24% of close contacts, i.e. health care-workers, household contacts, nursing home workers or residents[17]. Further, an observational study of Danish households showed SARS-CoV-2 transmission to 48% of household residents 14 days after the primary case tested positive[18].

Competitive swimming comprises less than 5% of DSF members but accounted for an estimated 39.4% of the three swimming activity categories’ pool activity hours during the period under study. When normalizing to pool activity hours the incidence rate analysis also suggests there was a higher rate of transmission during competitive swimming than during recreational swimming. This suggests that SARS-CoV-2 spreads easier in a competitive swimming setting than in recreational swimming. This is in line with the observation, that intense physical activity might increase susceptibility to upper respiratory infections[19]. However, this finding is contradictory to the results of a small cohort study, that training in competitive swimming does not change susceptibility to upper respiratory infections in general[20].

Yet, the apparent difference between competitive and recreational swimming might also be explained by more complete reporting from the competitive swimming activities. Though no statistically significant difference in sampling of risk episodes was found, swimming clubs in general have a better overview and more precise knowledge of competitive than recreational swimming activities, which could cause a sampling bias favoring reporting from competitive swimming. Also, it was not possible to control confounding factors like the age distribution between recreational and competitive swimming. Though, as individuals under 20 years of age are estimated to have half the susceptibility to SARS-CoV-2 infection than adults above this age[21], and most Danish competitive swimmers are below 20 years of age, susceptibility by age is not likely to explain the results.

The data suggest a high compliance to most of the 10 restrictions in the risk episodes during the pandemic. The highest compliance was to hand hygiene, which is higher than some previous research on behavior during the COVID-19 pandemic[22-24] and on level with other[25]. The risk assessment analysis failed to demonstrate a correlation between individual restrictions followed or the number of restrictions followed and the risk of transmission of SARS-CoV-2. Low statistical power suggest that this may have been due to low number of transmission episodes and a small sample in general. As we sampled the risk episodes in a comprehensive proportion of swimming clubs, a much larger sample might be difficult to achieve. In addition to lack of power, one should keep the risk of social desirability bias in self-reported data in mind when interpreting these results, as they may not mirror the true compliance (see limitations below).

A few reported risk episodes associated with activities in Danish swimming clubs were not included in the analysis since they included other activities than indoor swimming sessions, e.g. sauna after outdoor swimming and boxing training. Of special notice is a training camp lasting eight days, at which 23 swimmers from the same club were infected with SARS-CoV-2. Other clubs training in the same swimming pool in the same period did not report infected participants, and the exact time and source of transmission was never identified. Although transmission may have occurred during swimming activities at the training camp, this cannot be isolated from transmission associated with other activities during the training camp, such as sleeping in dormitories, dining and socializing together.

### Limitations

This study had several limitations. First, the retrospective design makes the study prone to recall bias. Second, some clubs reported to the survey based on logbooks of activities with notes of all risk episodes and results of follow-up tests, whereas other clubs may have collected the information while answering the questionnaire.

Some degree of recall bias may also be assigned to respondents’ hope for a certain outcome: Some respondents wrote as a final comment, that they looked forward to seeing the study results, as they were hoping for restrictions to be removed. This might have interfered with their recalling of compliance to individual restrictions at risk episodes. As noted above, the precise knowledge also may have varied depending on the respondents’ role in the club, which adds variability to the general recall bias. Only a minor fraction of respondents were trainers or instructors being present at the swimming pool during training and competition. Therefore, some respondents may have reported intended rather than actual compliance to some restrictions.

It was not possible to isolate the circumstances of risk episodes to be swimming activities exclusively. Risk episodes possibly involved contact between participants in the changing room, during shower and during transportation to and from the swimming pool premises. These activities may also differ between swimming activity categories. Also, as competitive swimmers generally are young people, they might attend the same school, e.g. dedicated classes for sport talents.

It is still debated how large a proportion of infected individuals are asymptomatic but able to transmit the virus to others[26]. A study showed 7.9% (6.6-8.8%) of Danish blood donors to be seropositive in week 6 in 2021 (8 - 14 February)[27]. At the same time, only 3.5% of the Danish population had been tested positive for SARS-CoV-2 at least once[16]. The discrepancy may indicate a large number of people not being tested even though tests during most of the pandemic were free an easily available in Denmark. As the identification of risk episodes in this study depended on the detection of infected participants through voluntary screening by publicly available PCR tests, the presence of infected but non-tested (likely asymptomatic) participants in swimming activities would not have been recorded. Consequently, the real number of risk episodes may therefore have been higher than the reported number. However, the good fit of data to the regional and national distribution of SARS-CoV-2 positive cases supports the assumption that the recorded risk episodes were a representative sample of the population of risk episodes. This also supports the data collection as valid to estimate the primary outcome.

To unify reporting and prevent recall bias of risk episodes, we defined a risk episode as “one infected person participating in swimming activities”, no matter how many swimming activities the person participated in. Accordingly, some infected subjects may have participated in more than one swimming activity before they were tested SARS-CoV-2 positive. The reporting of number of participants in risk episodes should partly compensate for the resulting underestimation of risk episodes, as this reporting enabled the respondent to report the total number of participants exposed to the presence of the SARS-CoV-2 positive participant defining the risk episode.

The reporting of transmission episodes was limited by the ability of the swimming clubs to ensure testing of all participants at a risk episode. According to widely communicated national guidelines, subjects in close contact with SARS-CoV-2 infected subjects should be tested. At all 162 risk episodes used to estimate transmission fractions and incidence rates, all participants were informed about these guidelines and asked to follow them. For the analyses, it was assumed that this instruction was followed, but the degree of compliance to the instruction cannot be quantified in this study.

No confounding factors were identified. As transmission was a rare event, the statistical analyses would have needed a larger sample to have enough power. However, as stated, the study included 82.7% of all members in DSF member clubs, so the potential for increasing sample size is limited.

## Conclusions

Based on replies from 172 Danish swimming clubs eight out of 162 risk episodes (4.9%) lead to infection of 23 participants, which constituted an estimated 1.0 (95% CI: 0.9;1.2)% of all participants in risk episodes. This corresponds to transmission of SARS-CoV-2 to 19.5 participants per 100,000 pool activity hours.

Compliance to 10 restrictions at risk episodes was between 69.9% and 98.1% with hand hygiene having highest compliance. No individual restrictions were found to change the risk of transmission, but this analysis had low statistical power.

## PERSPECTIVES

Controlled exposure experiments are very difficult to conduct using the SARS-CoV-2, but due to the unmatched surveillance of infections, the COVID-19 pandemic is a unique setting to investigate respiratory infections across different sports. As the most commonly reported infectious diseases in sports are viral and bacterial skin infections, this is a great chance of extending our knowledge[28].

The generalizability of the results is limited by the special circumstances during the restricted opening of Danish swimming activities in the second half of 2020. Nonetheless, the results provide a unique overview of risk episodes and transmission episodes in a large cohort taking part in swimming activities during the COVID-19 pandemic. These data are valuable as a foundation to design and implement restrictions on sports activities, seeking an optimal balance between costs and benefits in future pandemic or local epidemic scenarios.

## Supporting information

STROBE Statement

## Data Availability

The data that support the findings of this study are available on request from the corresponding author, SF. The data are not publicly available due to information that could compromise the participating swimming clubs' privacy.

## Data availability statement

The questionnaires in Danish and the data that support the findings of this study are available on request from the corresponding author, SF. The data are not publicly available due to information that could compromise research participants’ privacy (swimming clubs).

## Acknowledgements

The Danish Swimming Federation is thanked for collaborating in this study by distributing the questionnaire to its member clubs, sending out reminders and performing follow-up phone calls. The authors further would like to thank development consultant Lars Bo Larsen of the Danish Swimming Federation for sharing his expertise and knowledge on organized swimming activities in Denmark.

## Author contributions

MBT, AVC and SF conceived and designed the study. MBT collected data in collaboration with Danish Swimming Federation. MBT and SF analyzed the data. MBT prepared tables and figures. MBT, AVC and SF interpreted the data and wrote the manuscript, with MBT writing the first draft and AVC and SF revising during several rounds. MBT, AVC and SF all approved the final version and are responsible for the study according to the policy for research integrity, freedom of research and responsible conduct of research at Aarhus University.

